# Sharing positive changes made during COVID-19 national lockdown: a multi-method co-production study

**DOI:** 10.1101/2021.03.03.21252809

**Authors:** Lynn Williams, Bradley MacDonald, Lesley Rollins, Xanne Janssen, Leanne Fleming, Madeleine Grealy, Alison Kirk, David Young, Paul Flowers

## Abstract

**Objective:** A multi-method co-production study was designed to share psychosocial insights into the adoption of positive changes made during COVID-19 national lockdown in Scotland. We examined: i) the psychosocial patterning of positive changes, ii) the psychosocial processes by which positive change was realised, and worked with partner organizations to share our insights.

**Methods:** A sequential multi-method design included an online survey (n=2445) assessing positive changes in sleep and physical activity patterns, socio-demographics, mood, social support, coping, and resilience, with multivariate logistic regression analysis. We also employed interviews with a purposive diverse sub-sample of people self-reporting high levels of positive change (n=48) and used thematic analysis. Finally, partnership work translated insights into positive change-sharing targeted resources.

**Results:** The survey identified positive change was significantly patterned by age, gender and vulnerability to COVID-19. Higher positive reframing and higher active coping were associated with higher levels of cross-domain positive change. Higher symptoms of depression, planning, and self-distraction were associated with less cross-domain positive change. Thematic analysis showed the centrality of perceptions of time, opportunities to self-reflect and engage with the natural world, access support in diverse ways, actively build routine and purposefully build self-efficacy and a sense of control were key to initiating positive change. Our partner organizations focused on the rapid co-production of a series of online resources that shared study insights.

**Conclusions:** Our study, based around a salutogenic ethos and the constraints of COVID-19, sought to identify and share insights into achieving positive changes at a time of international crisis.

---

Globally COVID-19 national lockdowns have been the most significant public health interventions within living memory. While they have been successful in relation to their primary goal of reducing exponential transmission of COVID-19, they have also yielded a range of unintended consequences (see supplementary file 1 for a logic model describing a high-level overview of lockdown as a complex public health intervention). Many of these consequences have been pathogenic, particularly in relation to the worsening of mental health (O’Connor et al., 2020) and amplification of pre-existing health inequalities (Wright et al., 2020). As a counterpoint, here we take a novel salutogenic perspective examining and then sharing the positive changes people reported. Through our multi-method approach we examine the psychosocial patterning of these positive changes and the psychosocial processes by which positive change was realised. In addition, we report on how our community partners rapidly developed resources to share the study insights with people who could benefit from them. To our knowledge this study is the first in the world to focus first on a multi-method, salutogenic approach to COVID-19, and second to report on the rapid co-production of research-led resources intended to share insights into initiating positive change with people who may benefit.

Salutogenic approaches focus on the factors that support and promote health, particularly during stressful conditions (Antonovsky, 1979; Bauer et al., 2020). Within the context of COVID-19, applying this framework facilitates an examination of the positive adaptation and growth that may have been experienced by some individuals, including what we can learn about how and why some people have adapted and thrived during the national lockdown context and what might be amenable to change within the wider population. Salutogenesis can be realised through modifiable behaviors, personal resources, social capital or capabilities. It can include issues such as socioeconomic factors, social support, mood, coping and resilience, defined as “assets” which are important for achieving positive health outcomes.

The ability to adapt positively to COVID-19 may be linked to resilience, at both an individual and community level. It has been shown that when faced with stress and adversity some individuals are more psychologically resilient than others (Luthar & Cicchetti, 2001). Killgore at al. (2020) examined the factors that were associated with greater psychological resilience during the early stages of the nationwide lockdown in the United States. They found that resilience was higher for those who spent more time outside, exercised more, slept more and perceived better levels of social support. These findings highlight the importance of positive health-related behaviors in contributing to an individual’s psychological resilience. In addition, research has shown that resilience at the community level, including community cohesion and collective efficacy, can enable people to react positively to crises, such as natural disasters and epidemics (e.g. Alonge et al., 2019; Thornley et al., 2015). Bolstering community resilience, particularly in vulnerable communities, may aid the COVID-19 response and recovery process (Penkler et al., 2020).

Other factors linked to the successful adaptation to stress are social support and coping (Ong et al., 2006). Emerging evidence suggests an important role for these “assets” in our responses to COVID-19. For example, Fluharty and Fancourt (2020) found that those who were using avoidant coping styles included those with lower socioeconomic status, higher levels of loneliness and those experiencing COVID-19 related adverse events. These groups have also been identified as experiencing poorer mental health during the pandemic, suggesting that coping strategies can influence how effectively we manage the stress of the pandemic. In contrast, Gori et al. (2020) found that the use of approach coping (including planning and active coping) was associated with lower levels of COVID-19 stress.

In addition to the role of psychological factors in enabling positive adaptation to COVID-19, social factors are influential. Recent evidence has suggested a social patterning of positive changes experienced during COVID-19 national lockdown with findings revealing differences in the amount of positive changes that different sub-populations have experienced (Cornell et al., 2020; Williams et al., 2021). Specifically, those reporting higher levels of positive change were female, from younger age groups, married or living with their partner, and in better health. These results suggest that some groups were better able than others to take advantage of national lockdown as an unexpected catalyst for positive change.

Both Williams et al. (2021) and Cornell et al. (2020) also report on the types of positive changes that people experienced during the pandemic. The positive changes reported by Williams et al. (2021) in their survey of Scottish adults included: being more appreciative of things usually taken for granted, spending more time doing enjoyable things, having more time in nature or being outdoors, paying more attention to personal health, increasing physical activity, and having more quality time with partner or spouse. In their study of Australian adults, Cornell et al. (2020) found three main positive changes: the opportunity to spend more time with family, appreciating having more flexibility in work arrangements, and enjoying a less busy lifestyle.

Positive changes have also been observed in health-related behaviors, with Janssen et al. (2020) reporting that although levels of sedentary behavior increased during lockdown, levels of moderate to vigorous activity increased and participants reported sleeping longer at night. It was also noted that participants who changed one behavior positively were also more likely to report a positive change in another demonstrating an important clustering in positive behavior change. Within the survey component of the current study, we further investigate this clustering of positive behavior change by focusing our analysis on a cross-domain measure of positive behavioral change, incorporating sedentary behavior, physical activity, and sleep. Doing so allows us to examine the factors associated with positive change beyond those associated with one specific outcome (e.g. physical activity alone) and also allows us to integrate the findings with the qualitative study where participants tell us about positive change across multiple domains.

In the current multi-method study, we investigate cross-domain salutogenic outcomes experienced through national lockdown. We examine the psychosocial patterning of these positive changes and the psychosocial processes by which positive change was realised. In addition, we outline the process by which we worked with partner organizations to share our insights and co-produce positive change-sharing, targeted resources suitable for each organization’s key audience/community base. Consequently, we have three key research questions (RQs):

RQ1. Which psychosocial variables were associated with making cross-domain positive change?
RQ2. What were the psychosocial processes by which positive change was initiated?
RQ3. How can we rapidly share this learning with partner organizations to co-produce positive change-sharing targeted resources?

## Method

The sequential multi-method design involved a quantitative cross-sectional online survey and qualitative in-depth interviews (reported according to the Mixed Methods Article Reporting Standards (MMARS; Levitt et al., 2018). Subsequently findings were integrated and then translated into simple resources to enable our partner organizations to develop tailored resources that were appropriate for their audiences. All materials and procedures were approved by the Ethics Committee of the University of Strathclyde, Scotland, UK (Ref 61/05/05/2020 A Williams) and all participants provided informed consent.

### Quantitative study

The survey sample comprised 2445 participants (participant characteristics are shown in Table 1). Data collection took place from 20 May 2020 to 12 June 2020, spanning the 9th to the 12th week of the first national lockdown in Scotland. This period was part of the first UK-wide lockdown that commenced in March 2020. People were advised to ‘stay at home’ and work from home wherever possible, only leaving home for essential purposes. These essential purposes included food shopping, medical appointments or to provide care for a vulnerable person. People were also allowed to leave home for one form of exercise each day. The target population of the survey was adults, aged 18 years or older, currently residing in Scotland, who were interested in sharing their experience of positive change. Participants were primarily recruited through social media advertisements on Facebook and Twitter, which directed participants to the online survey on Qualtrics.

**Table 1.** Descriptive sample statistics for socio-demographic and health variables

| Variable |  | N | % |
| --- | --- | --- | --- |
| Age | 18-24 | 236 | 9.7 |
|  | 25-34 | 411 | 16.9 |
|  | 35-49 | 638 | 26.2 |
|  | 50-64 | 838 | 34.4 |
|  | 65+ | 312 | 12.8 |
| Gender | Female | 1956 | 81.5 |
|  | Male | 451 | 18.5 |
| Household income | <£16 000 | 238 | 10.9 |
|  | £16 000-£29 999 | 431 | 19.8 |
|  | £30 000-£59 999 | 876 | 40.3 |
|  | £60 000- £89 999 | 407 | 18.7 |
|  | £ 90 000+ | 224 | 10.3 |
| Employment status | Employed | 1649 | 67.6 |
|  | Inactive (e.g. retired, furloughed) | 754 | 30.9 |
|  | Unemployed | 37 | 1.5 |
| Highest education level | High School | 258 | 10.8 |
|  | College | 360 | 15.1 |
|  | Undergraduate | 723 | 30.4 |
|  | Postgraduate | 1041 | 43.7 |
| Ethnicity | White | 2361 | 97.4 |
|  | BAME | 64 | 2.6 |
| Relationship status | Single | 440 | 18.2 |
|  | Married or living with partner | 1601 | 66.2 |
|  | Have a partner but not living together | 187 | 7.7 |
|  | Separated /divorced or widowed | 191 | 7.9 |
| Health | Very poor | 6 | 0.2 |
|  | Poor | 43 | 1.8 |
|  | Fair | 340 | 14.0 |
|  | Good | 1113 | 45.7 |
|  | Very good | 932 | 38.3 |
| High-risk | Yes | 370 | 15.2 |
|  | No | 2070 | 84.8 |

### Measures

#### Socio-demographic characteristics

included age, gender, relationship status, ethnicity, education, income, and employment status. Participants also self-reported on their perceived health and whether they were high risk/shielding for COVID-19. Sample characteristics are shown in Table 1.

#### Behavioral outcomes

physical activity and sedentary behavior were assessed by the short form of the International Physical Activity Questionnaire (Craig et al., 2003) to derive duration of time spent sitting, walking, and in moderate-to-vigorous physical activity (MVPA). Sleep duration was assessed by self-reported number of hours sleep per night. Participants completed these behavioral measures to reflect a normal week pre-lockdown and their current behavior (i.e. during national lockdown).

#### Psychological measures

Social support was assessed by the Brief Perceived Social Support Questionnaire (Lin, Hirschfled & Margraf, 2018). The Brief COPE inventory (Carver et al., 1997) was used to assess the seven coping strategies of positive reframing, acceptance, planning, humour, active coping, emotional support, self-distraction, and instrumental support. The resilience dimensions of personal competency and social resources were assessed by the Resilience Scale for Adults (Friborg et al., 2005). Symptoms of depression and anxiety were assessed by the 4-item Patient Health Questionnaire (Kroenke et al., 2009).

#### Statistical analysis

Following the process of Janssen et al. (2020) we categorised the changes in sleep, sitting, walking and MVPA that participants had experienced from pre-lockdown to lockdown as either ‘positive change’, ‘negative change’, ‘no change’ and ‘already optimal’ based on previously reported categorizations for levels of physical activity (Ekelund et al., 2016), sitting (Stamatakis et al., 2019) and sleep (the National Sleep Foundation’s Sleep Duration Recommendations). As our interest is in cross-domain positive change we created an additive cross-domain positive change score as primary dependent variable. This was the % of possible positive changes achieved on the four behavioral domains (sitting, walking, MVPA, sleep). Utilising a % score allowed us to factor in domains for which participants were already scoring optimally with no positive change possible. For example, if someone was already sleeping optimally pre-lockdown no further positive change could be recorded and therefore the % score was calculated as: number of positive changes divided by three instead of four. We then created a dichotomous variable for the purposes of logistic regression analysis reflecting participants who had made 50% positive change or more (n=926) vs. participants who had made less than 50% positive change (n=1519). We made the decision to create this new dichotomous outcome variable instead of using the original % scores because of the complexity of the underlying % values which resulted in seven different % possibilities, which were not equally spaced (see supplementary file 2 for the distribution of these scores). Creating the new dichotomous variable therefore gives us a more interpretable outcome for the logistic regression analysis. Univariate and multivariate binary logistic regression analyses were carried out to determine the factors associated with cross-domain positive change. All analyses were conducted using IBM SPSS Statistics (version 25) at 5% significant levels.

#### Qualitative study

A purposive sub-sample of survey respondents reporting high levels of positive change was identified. Within this larger sample, using a sampling matrix, we sought to recruit a purposive sample in relation to representing those from diverse ethnic backgrounds, an equal span of gender, a range of educational attainment, income and diverse relationship status. Of 118 people approached 48 took part in interviews (see Table 2 for the sociodemographic characteristics of the interview participants). Interviews were conducted by the research team after bespoke training had ensured shared approaches to data collection drawing on interpretative phenomenological analysis (Smith, Flowers and Larkin, 2009). A topic guide was used within the interviews to guide the interaction; this covered the broad arc of people’s COVID-19 experiences from early on within the pandemic, through initiating and then maintaining positive changes. Throughout, participant disclosures were followed by tailored questions concerning thoughts and feelings relating to their experiences. One interviewer (BM) conducted the majority of interviews (n=39). Remote semi-structured interviews took place between 12 June and 24 September 2020 and were approximately 50 minutes in length. Interviews took place at a variety of times and in a range of modalities according to participant preference, and included platforms such as Zoom, Skype and telephone. Audio recordings were collected using dedicated voice recording devices only.

**Table 2.** Socio-demographic characteristics of the interview participants

| Variable |  | N | % |
| --- | --- | --- | --- |
| Age | 18-24 | 6 | 12.5 |
|  | 25-34 | 10 | 20.8 |
|  | 35-49 | 9 | 18.7 |
|  | 50-64 | 18 | 37.5 |
|  | 65+ | 5 | 10.5 |
| Gender | Female | 24 | 50 |
|  | Male | 23 | 48 |
|  | Transgender (female) | 1 | 2 |
| Household income | <£16 000 | 6 | 12.5 |
|  | £16 000-£29 999 | 11 | 22.9 |
|  | £30 000-£59 999 | 15 | 31.3 |
|  | £60 000- £89 999 | 7 | 14.6 |
|  | £ 90 000+ | 4 | 8.3 |
|  | Prefer not to say | 5 | 10.4 |
| Employment status | Employed | 32 | 66.7 |
|  | Inactive (e.g. retired, furloughed) | 16 | 33.3 |
|  | Unemployed | 0 | 0 |
| Highest education level | No qualifications | 2 | 4.2 |
|  | High School | 16 | 33.3 |
|  | College | 3 | 6.3 |
|  | Undergraduate | 8 | 16.6 |
|  | Postgraduate | 19 | 39.6 |
| Ethnicity | White | 26 | 54.2 |
|  | BAME | 22 | 45.8 |
| Relationship status | Single | 10 | 20.8 |
|  | Married or living with partner | 28 | 58.3 |
|  | Have a partner but not living together | 4 | 8.3 |
|  | Separated /divorced or widowed | 4 | 8.3 |
|  | Other | 1 | 2.1 |
| Health | Very poor | 0 | 0 |
|  | Poor | 2 | 4.2 |
|  | Fair | 6 | 12.5 |
|  | Good | 22 | 45.8 |
|  | Very good | 18 | 37.5 |
| High-risk | Yes | 4 | 8.3 |
|  | No | 44 | 91.7 |

Transcripts were uploaded to coding software program Nvivo. Drawing on Braun and Clarke’s (2006) approach to thematic analysis, across the project initial inductive open coding (n=27) was followed by more deductive coding (n=21) as the analytic framework developed and was reified over time. Initial analysis was conducted by one researcher (BM) under the supervision of a senior qualitative research expert (PF). Supervision consisted of focussed discussions on data interpretation, thematic coherence, and the inter-relationships between the thematic entities.

#### Integration of findings

To integrate the findings of the two contributing studies, their key results were firstly entered into a single table. Drawing on the constant comparative method (Glaser and Strauss, 1967) two researchers (LW and PF) independently analysed the key results, focusing on areas of similarity and difference and generating interpretative statements of the overall meaning of the two contributing studies. Subsequently they met to share their interpretations and agree a consensus position. Their work was then audited by the wider CATALYST team to ensure its validity, coherence and substantiation.

#### Partnership work

We engaged in partnership work with three charity organizations with the aim of rapidly developing public-facing resources that could share insights from the project with those who might benefit the most. Community partner organizations were selected on the basis of our early analyses of the social patterning of positive change (Williams et al., 2021). We had found that men had experienced less positive change so we approached an organisation specialising in the promotion of men’s mental health (Mind the Men). We had also found that older people were less likely to report positive changes so we approached an organisation that focussed on older people’s wellbeing (Scottish Older People’s Assembly). To complement these population-specific organizations we chose to approach one organisation with a particular behavioral focus on promoting physical activity (Actify). To capitalise on our community partner organizations’ expertise at working with their audiences/client base and their unique knowledge of how best to effectively communicate with them, we took an approach of translating our findings into a short report that provided a lay summary of the key findings from the qualitative components, provided ‘top tips’ for initiating positive change, and gave exemplar quotes from the interview transcripts (see supplementary file 3). More dialogue then followed.

We took a flexible approach with our partners about what they wished to do with our findings and enabled them to choose what and how to develop resources and in which modalities. The final resources developed by our partners included infographics, short films, and downloadable materials shared on the websites and social media platforms of the different organizations. All materials were targeted at awareness raising and positive change-sharing in order to facilitate the initiation of positive health change among those who may benefit.

## Results

### Quantitative study

As shown in Table 1, there was a good age distribution among participants from 18-24 (9.7%), 25-34 (16.9%), 35-49 (26.2%), 50-64 (34.4%) to 65+ (12.8%). The majority of the sample were female (81.5%). Most participants were employed (67.6%) and educated to university level (74.1%). In terms of self-reported health, 15.2% identified that they were at high-risk of COVID-19 or were “shielding” based on underlying health conditions or age.

### Factors associated with cross-domain positive change

Univariate analyses showed that ethnicity (p=0.105), education (p=0.506) and income (p=0.072) were not associated with the extent of cross-domain positive changes made and were therefore not considered in a multivariate model. The following factors were significant and were therefore included in a multivariate logistic regression analyses using stepwise selection: age, gender, relationship status, employment status, self-reported health, being in a high-risk group, social support, anxiety, depression, both of the resilience variables and all of the coping variables (full details of the outcome from the univariate analyses can be found in supplementary file 4).

In the final multivariate model eight factors emerged as significant predictors: these were age, gender, high risk status, symptoms of depression, positive reframing, planning, active coping, and self-distraction. When considering the coefficients (see Table 3), those aged 25-34, 35-49 and 50-64 were all more likely to report making ≥50% cross-domain positive changes compared to those 65+. Those who were female were more likely to report making ≥50% cross-domain positive changes compared to males. Those who were shielding were less likely to have reported positive cross-domain change. Higher levels of positive reframing and higher active coping were associated with being more likely to have made ≥50% positive cross-domain changes. Higher symptoms of depression, and higher levels of planning, and self-distraction were associated with being less likely to have made positive changes across domains.

**Table 3.** Multivariate analysis of socio-demographic and psychological factors predicting ≥50% positive change

| **Variable** | **p-value** | **Comparison** | **Coefficient** | **95% CI** | **p-value** |
| --- | --- | --- | --- | --- | --- |
| Age | .007 | 18-24 vs. 65+ | 1.37 | .89-2.16 | .158 |
|  |  | 25-34 vs. 65+ | 1.65 | 1.13-2.39 | .009 |
|  |  | 35-49 vs. 65+ | 1.87 | 1.33-2.63 | <.001 |
|  |  | 50-64 vs. 65+ | 1.64 | 1.18-2.28 | .003 |
| Gender | .016 | Female vs. Male | 1.35 | 1.06-1.72 | - |
| High risk/Shielding | .026 | Yes vs. No | .722 | .54-.96 | - |
| Depression | <.001 | - | .846 | .79-.90 | - |
| Positive reframing | <.001 | - | 1.32 | 1.16-1.50 | - |
| Planning | .016 | - | .789 | .65-.96 | - |
| Active coping | .013 | - | 1.28 | 1.05-1.56 | - |
| Self-distraction | <.001 | - | .819 | .73-.92 | - |

### Qualitative study

The inductive thematic analysis highlighted six inter-related themes which reflect key psychological processes reported by the participants. Although the themes are presented in a linear fashion it is important to remember that they are inter-related, as shown in Figure 1.

**Figure 1.**
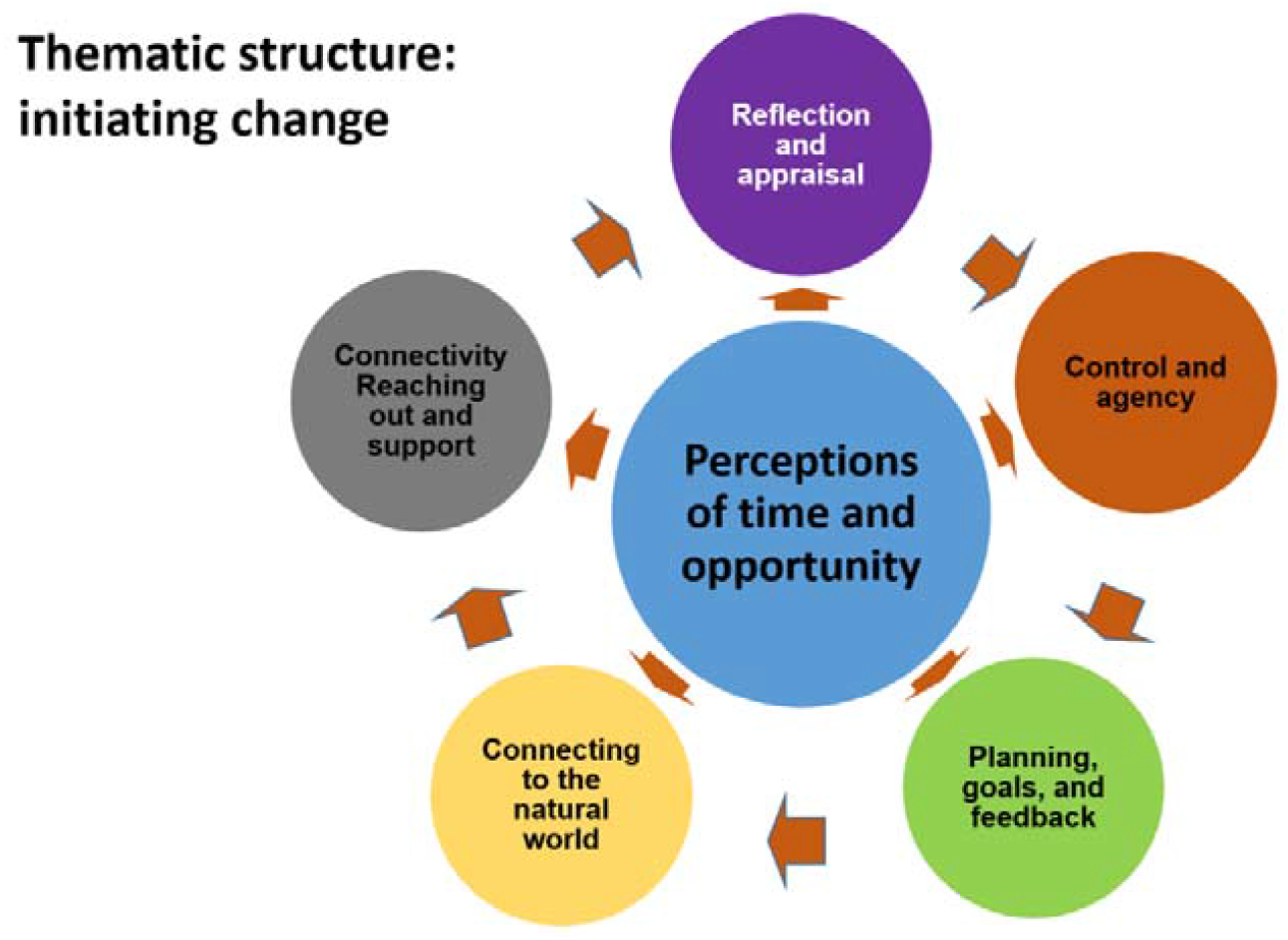
Depiction of the inter-relationship of key themes

### Time and opportunity: *‘the construct of time has changed over the last couple of months’* (IP5)

Participants talked about how lockdown had removed many of the life’s routines and associated demands. Regular time commitments such as commuting, transporting children, many forms of socialising, or caring duties were removed. For some, these new time opportunities were quickly filled with other demands such as child care and home schooling, but for many participants it was as if the momentum of pre-COVID life was lost and the passage of time distorted, giving them a new clarity about time as a resource. They were able to think about, and enact, new ways of restructuring their lives in ways that worked positively for them:-

> Well, I suppose my life’s slowed down massively. Like, we’re really busy with family, we’ve got two kids, and you know, like, basically during the week it’ll be work and then there’ll be, kids’ll be at clubs every night and some nights won’t be sitting down ‘til eight o’clock at night, you’re absolutely knackered. I suppose having people over all the time – like, the kids have friends over all the time, and maybe being forced to stop it made me realise that we needed to slow the pace down massively because it wasnae actually enjoyable before. Even the kids I think much preferred a coupla more nights at home rather than being at clubs all the time. (IP42)

> With lockdown, you know, you sort of like lose the whole structure. So you do… at least, I realised that I do have a choice to dictate what I want to do in the morning, you know, or what time I want to end the day. So it could, I could just… I could choose to, you know, call it an early day at two o’clock, or I could, you know, push all the way until eight, nine pm and that’s fine. (IP1)

In summary, this theme has shown how lockdown brought an apparent slower pace of life which facilitated an evaluation of *how* they wanted to manage time and live their lives more positively where possible.

### Reflection and appraisal: *‘it gave me time to reflect on kind of what’s important in my life’*

As indicated above many participants reported that lockdown facilitated a ‘stepping back’ and appraisal of their lives and the way they were living and their life priorities. Participants talked of lockdown as meaning that they were ‘*escaping the noise’* (IP35) and were able to reflect on their lives in new ways. As one participant said, such reflections were *‘quite fundamental to – to making change you actually want to, like, see’*. (IP50). The extracts below illustrate the importance of this reflection and appraisal:-

> I suppose it gave me time to reflect on kind of what’s important in my life.[…..] So it was kinda reflecting, you know, reflecting on things that were important and what you do and why you do them, if you know what I mean? (IP41)

Appraisal and reflection were important as they enabled participants to generate solutions to on-going problems such as key aspects of their relationships, or developing new solutions to issues such as sedentary behavior or diet. For example:-

> I believe for the first time in my life – and I think my wife has experienced the same thing – we’ve had the kind of psychological breathing room to kind of understand ourselves and our difficulties. So whether those are mental health difficulties or for me a lot of it’s been difficulties in kind of conceptualising ideas. A lot of that has now kind of all come together with this new-found kind of freedom […….] And I think that’s really what has – if you’re in the right mental space – has given us the capability to look at our situation in quite a different way. (IP35)

In summary this theme has shown how the lockdown restrictions provided distinct opportunities for some participants to reflect and appraise key aspects of their lives. This was pivotal to understanding the adoption of various positive changes.

### The limits of control and agency: *‘focus on what you can do, what you can control, and what you can’t control’*

This theme highlights the importance of perceived control in shaping the initiation of positive change. As global events, legislation, government policy and news reporting continually highlighted a lack of personal control, many participants had initially felt deeply distressed. However, the identification of the distinction between things that were possible to change (e.g. physical activity, immediate environment and work tasks) and things that were not, was centrally important for this sample of people reporting high levels of positive change:

> I think when you’re just forced into like, you’ve got more, I just thought, ‘Well life’s slowed down considerably,’ and we had a lot more time to think about things and I suppose I wasn’t a very good, it just felt like my life had been put on hold and I didn’t really have control over anything anymore. And there were only a couple of things that you really had control over. So I just decided to control the few things that I could. Just before we went into lockdown I was actually looking for another job and now kinda got put on hold because of this and I was getting a bit frustrated so I thought, ‘Well this is the one thing that I can have some control over is how much I can walk, how long I can go for a walk. Or, you know, what kind of exercise I can do at home, like…’ I think it was one of the few things, so I just, yeah, decided to try and make some small changes. (IP47)

For others, the locus of what they perceived as changeable related to their immediate environment:-

> I would also say, you know, what has been… you know, in my kinda darkest days, sometimes just breaking it down to basics is helpful. So if I’m, if I’m having a really crappy day and I’m not feeling good at all, I will just clean and hoover, and there’s two elements to this: one is, you know, I’m gonna have to be in this space, I want it to be nice and pleasant and tidy and clean; the other is, you know, even if I feel like completely powerless and completely out of control in terms of what’s happening in the world, I can still control how clean this room is. And that gives me that sense of, you know, this is my world, I can run my world the way I want to run my world. The rest of the world we’ll get to in a minute, but for now, let’s hoover the rug, you know? (IP25)

In summary this theme has highlighted how perceptions of a loss of control due to COVID-19 were often effectively countered by the identification of pragmatic locus of control, which in turn enhanced feelings of agency and self-efficacy.

### Planning, goals, and feedback: *‘I decided to get a routine’*

This theme highlights how processes of planning, goal setting and developing new habits were important in relation to positive change. Many of the participants highlighted that these processes were vitally important to them. The extracts below show how the extra time afforded by lockdown was important in thinking about goals and making the planning of activities associated with positive change more realistic.

> Yeah, well, ‘cause people always have goals, but they [cuts out] can be quite vague. So during lockdown when you had more time to think about it, you’re like ‘Well, I’m not’ – I had so much less to do, why not just get some paper out, think about what I want, and then you can start creating an actual path towards that. (IP50)

Other participants talked about the importance of planning *per se* in enabling positive changes to occur:-

> Just keep – you know, have a kinda ‘to-do’ list every day, couple o’ jobs on it for yourself, your own sanity. Keep yourself busy. Like, make plans for around the house. Like, “Tomorrow I’m gonna paint the fence,” or whatever. Or go a walk to – or even, like, we did things wi’ the kids like scavenger hunts. But it would be like, insect scavenger hunts. You’d go a walk with a wee sheet and they would have to tick off what animals they found. (IP42)

In summary, this theme highlighted that establishing routines through planning and achievable goal setting were important catalysts to initiating positive changes during lockdown. Some participants charted their changes in diaries and others sought out and discussed their achievement with others.

### Connecting to the natural world: *‘going out watching the flowers coming out’*

This theme shows that during lockdown participants felt that connecting to and exploring the natural world, helped them to be more positive. The extract below introduces this theme and sharply contrasts the pre-COVID world with the experience of lockdown:-

> Normally, the media and everyone marketing tells us that to feel happy, we need to buy stuff or travel or buy [cuts out] and that will meet the sense emptiness or incompleteness that we have. Not emptiness, I think incompleteness. And that, of course, isn’t an option at the moment. Well, maybe one could buy stuff, but it’s not quite the same. So I hope, it’s certainly been my experience, I’ve, and my family, we’ve been rediscovering simple pleasures like watching the blue tits in the bird box. [Laugh] or a really obvious, not obvious, but vivid example. We have rhubarb in the garden. It just grows there. For the last twenty years, but this year, I have noticed it unfurling its leaves more. (IP22)

Unlike the previous themes here there is little sense of active thought processes or deliberate cognition. In contrast to those reflective, agentic ways of achieving positive changes, this theme captures how for many participants being in the natural world was simply cathartic. There was a passive pleasure in engaging with the natural world. The extracts below stress this sensory quality:

> at one point I got really far, and it was just like quiet and it was a really nice day, and, yeah, it was just… it sounded like the sea a little bit, there was like seagulls that day, and it smelled like the sea as well, a little bit. So that was quite nice. I think that was like my favourite run, it was one of the longest as well. (IP18)

In summary this theme has highlighted how connection to and engagement with the natural world was positive for many participants. Of note, this was often described as a passive process of being immersed within the natural world and of taking simple sensual pleasures from watching birds, seeing flowers open and change, or smelling the sea or hearing the wind.

### Connectivity, reaching out and support: *‘I was doin’ it for me but I was also doin’ it to help other people’*

This theme shows how positive change was achieved through reaching out to others for support, or providing support to others. Both routes were associated with positive changes to health and wellbeing. Much but not all of this support was digitally mediated. Several participants talked of using Instagram and Facebook for the first time, being connected to others through ‘likes’ and shared interests, ‘***little kind of sharing a day by day’ (IP14)***. Others used digital connectivity to form new social networks or to reinforce existing ones, to engage in new activities or reengage with old (ballets, choirs, watching online performances), to be part of activism and community engagement and to take an active role in fundraising. For some digital connectivity made social support in local neighbourhoods real:-

> it’s through social media. So it’s Facebook Messenger or text messages. So we tend to communicate with each other like that, so we’ll text in the morning and say, “Look, we’re off to Sainsbury’s, does anybody need anything?” And we’ll get them and then we drop the shopping off outside the door and stand back and you have that little blether for five minutes while they take the shopping in and it’s just really nice ‘cause you actually get to see each other and it’s really nice ‘cause you’re kind of looking for the text in the morning to see who’s doing what and if anybody’s going anywhere. And it means, and it’s just simple things like making sure nobody runs out of bread or milk and stuff and it’s been—just been really nice and I feel that we’ve really, really got to know our neighbours better than we ever thought we would because normally, you know, you’re like ships in the night. (IP44)

For some new communities were created:-

> I kind of realised that, you know, I wasn’t the only one struggling with isolation and loneliness because I did realise that there were other, you know, students who… on campus who were pretty much staying alone, too, because for whatever reason… you know, some of them couldn’t go back to their home country because the borders were closing kind of thing, and I realised that, oh, so I’m not the only one that’s stuck in this or, you know, facing this set of emotions, and actually we kind of like… after a while, we actually formed our own, like, you know, WhatsApp group, ‘cause, you know, we kinda like grouped together. You know, we would band together to do like quiz nights on Zoom, you know, share virtual recipes and things like that, ‘cause… you know, it’s kinda like a support group, you know, where we’d kind of like help each other get through the week, you know, and before the… before we knew it, we were like, oh, we’re done with April, you know, we’re done with May, and we were like, oh, okay, we’re in the middle of June now, and yeah. (Laughs) Yeah, yeah. (IP1)

In summary this theme has shown how important connecting with others was for making positive changes during lockdown. The digital media were key to enabling new connections to form and providing a means to meeting and engaging with others in the absence of risks associated with COVID-19 transmission.

## Discussion

The current study took a novel salutogenic perspective to examine and then share the positive changes people reported during COVID-19. Through our multi-method approach we examined the psychosocial patterning of these cross-domain positive changes and the psychosocial processes by which positive change was realised. The integration of the key findings from the quantitative and qualitative components is shown in Table 4. These findings highlight that enabling people to reframe the lockdowns, and particularly how they think about time, may be central to initiating positive change. They also suggest that taking active steps to adapt one’s life through proactive self-reflection and appraisal was key to recognising opportunities to make positive change. Our findings also suggested that re-asserting control through manageable goals was important. While the pandemic brought about a loss of control for many aspects of our lives, those that concentrated on what they could control, and plan for, in their lives day-to-day were able to make positive changes. The data also suggested that self-distraction via engaging with the natural world was important in making positive changes, as was social connectedness.

**Table 4.**
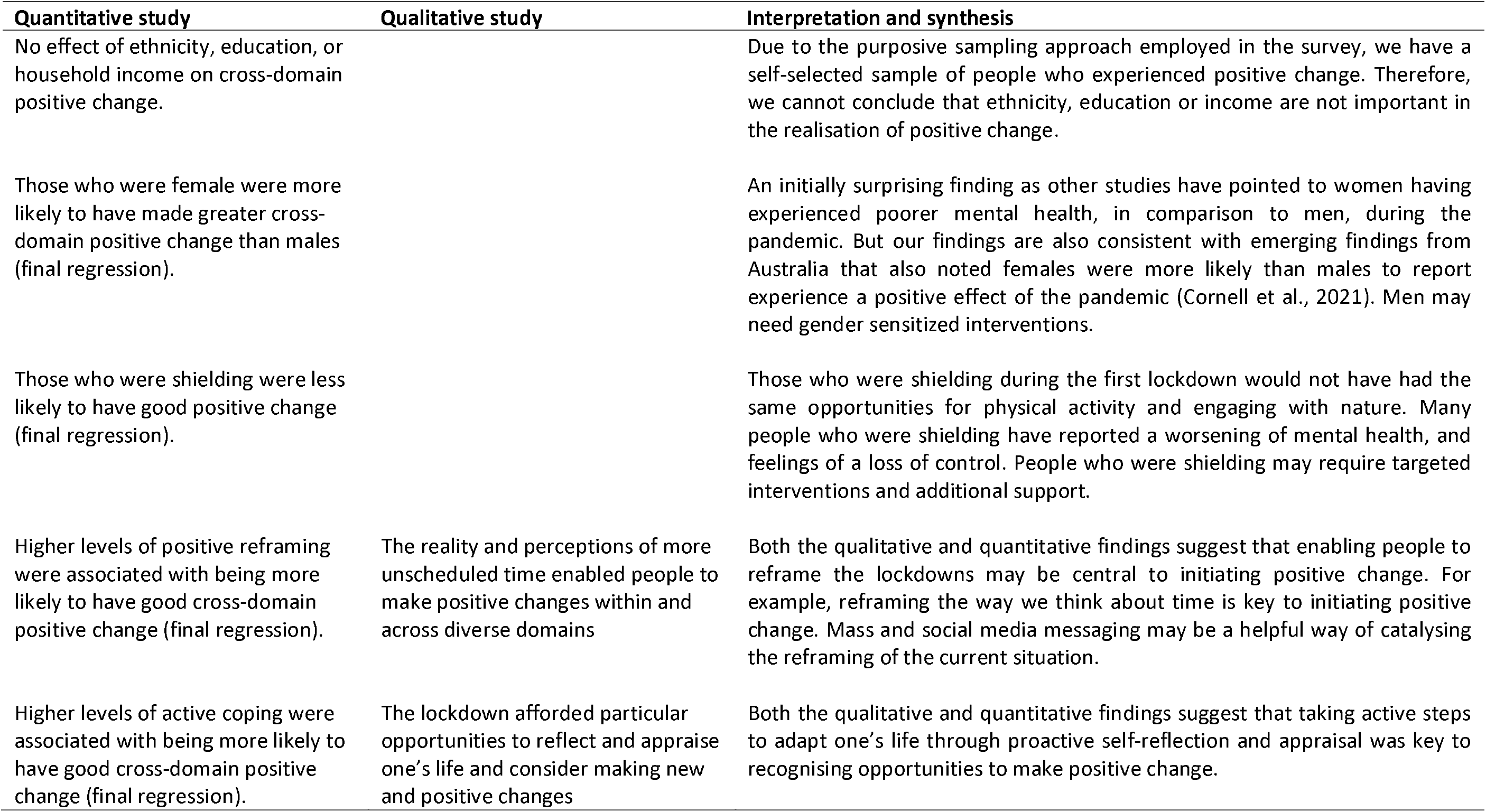
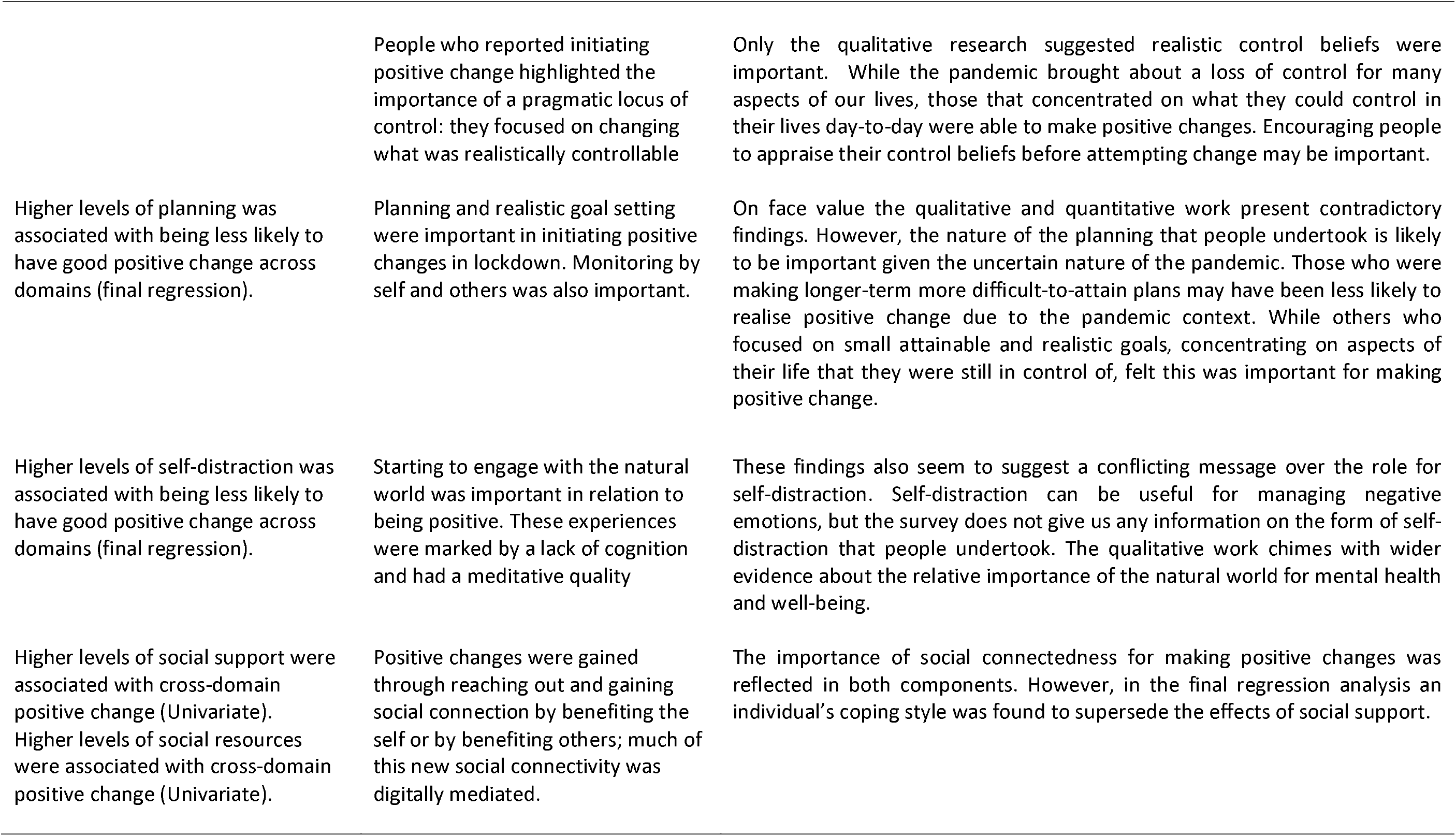
Integration of the main findings across the quantitative and qualitative studies

| Quantitative study | Qualitative study | Interpretation and synthesis |
| --- | --- | --- |
| No effect of ethnicity, education, or household income on cross-domain positive change. |  | Due to the purposive sampling approach employed in the survey, we have a self-selected sample of people who experienced positive change. Therefore, we cannot conclude that ethnicity, education or income are not important in the realisation of positive change. |
| Those who were female were more likely to have made greater cross-domain positive change than males (final regression). |  | An initially surprising finding as other studies have pointed to women having experienced poorer mental health, in comparison to men, during the pandemic. But our findings are also consistent with emerging findings from Australia that also noted females were more likely than males to report experience a positive effect of the pandemic (Cornell et al., 2021). Men may need gender sensitized interventions. |
| Those who were shielding were less likely to have good positive change (final regression). |  | Those who were shielding during the first lockdown would not have had the same opportunities for physical activity and engaging with nature. Many people who were shielding have reported a worsening of mental health, and feelings of a loss of control. People who were shielding may require targeted interventions and additional support. |
| Higher levels of positive reframing were associated with being more likely to have good cross-domain positive change (final regression). | The reality and perceptions of more unscheduled time enabled people to make positive changes within and across diverse domains | Both the qualitative and quantitative findings suggest that enabling people to reframe the lockdowns may be central to initiating positive change. For example, reframing the way we think about time is key to initiating positive change. Mass and social media messaging may be a helpful way of catalysing the reframing of the current situation. |
| Higher levels of active coping were associated with being more likely to have good cross-domain positive change (final regression). | The lockdown afforded particular opportunities to reflect and appraise one's life and consider making new and positive changes | Both the qualitative and quantitative findings suggest that taking active steps to adapt one's life through proactive self-reflection and appraisal was key to recognising opportunities to make positive change. |
|  | People who reported initiating positive change highlighted the importance of a pragmatic locus of control: they focused on changing what was realistically controllable | Only the qualitative research suggested realistic control beliefs were important. While the pandemic brought about a loss of control for many aspects of our lives, those that concentrated on what they could control in their lives day-to-day were able to make positive changes. Encouraging people to appraise their control beliefs before attempting change may be important. |
| Higher levels of planning was associated with being less likely to have good positive change across domains (final regression). | Planning and realistic goal setting were important in initiating positive changes in lockdown. Monitoring by self and others was also important. | On face value the qualitative and quantitative work present contradictory findings. However, the nature of the planning that people undertook is likely to be important given the uncertain nature of the pandemic. Those who were making longer-term more difficult-to-attain plans may have been less likely to realise positive change due to the pandemic context. While others who focused on small attainable and realistic goals, concentrating on aspects of their life that they were still in control of, felt this was important for making positive change. |
| Higher levels of self-distraction was associated with being less likely to have good positive change across domains (final regression). | Starting to engage with the natural world was important in relation to being positive. These experiences were marked by a lack of cognition and had a meditative quality | These findings also seem to suggest a conflicting message over the role for self-distraction. Self-distraction can be useful for managing negative emotions, but the survey does not give us any information on the form of self-distraction that people undertook. The qualitative work chimes with wider evidence about the relative importance of the natural world for mental health and well-being. |
| Higher levels of social support were associated with cross-domain positive change (Univariate). Higher levels of social resources were associated with cross-domain positive change (Univariate). | Positive changes were gained through reaching out and gaining social connection by benefiting the self or by benefiting others; much of this new social connectivity was digitally mediated. | The importance of social connectedness for making positive changes was reflected in both components. However, in the final regression analysis an individual's coping style was found to supersede the effects of social support. |

Many of the findings raise longer term questions about COVID-19 recovery and what people want for themselves, their families, their workplaces and communities. In particular, time and opportunity were found to be central and provided a gateway to the realisation of positive change. Participants noted that many of the routines and demands of pre-COVID life (often linked to time poverty) had been removed, and there was a perception by many that they now had more time available. People appreciated the slower pace of life that this afforded them, and it also facilitated a stepping back for many people, allowing them to appraise and reflect upon issues in their lives and reassess their priorities.

Our survey results also suggest that some groups experienced more cross-domain positive change than others. Amongst this sample of people recruited because they had experienced some positive change, older adults in particular, along with those who were ‘shielding’, males, and those with lower mood, made fewer positive changes. Previous work has also highlighted the social patterning of positive changes during COVID-19 (Cornell et al., 2020; Williams et al., 2021). The findings suggest that these groups, as well as other vulnerable and marginalised groups who have suffered the most through the pandemic, may require targeted interventions (e.g. age or gender sensitized) and additional support to enable them to make positive changes and reduce the harms caused by the pandemic.

A further aim of the study was to rapidly share our findings with partner organizations to co-produce positive change-sharing targeted resources. The final resources developed included short films, animations, and infographics (accessible via https://www.actify.org.uk/catalyst). These materials were shared on the websites and social media platforms of the different organizations. All materials were targeted at awareness raising and positive change-sharing in order to facilitate the initiation of positive health change among those who may benefit. These materials represent a novel and initial attempt at salutogenic intervention development within the context of COVID-19. In terms of the post-pandemic recovery phase, the scaling-up of these intervention resources and further development of systemic salutogenic-based interventions may be useful for enabling communities to change positively and build resilience.

Our study also speaks to how we should operationalise and theorise salutogenisis. Although these focal points were not a primary concern of the work it is interesting to reflect on what we have learned and the uniqueness of conducting health psychology research within a rapidly unfolding pandemic. Our decision to focus on learning from those who experienced positive change, and then share these insights with people who could potentially benefit from it, did shape our overall design and analytic foci. For example, we did not attempt to recruit a nationally representative sample for either the quantitative or the qualitative research. In relation to theory, although we began with a broad interest in salutogenisis we did not operationalise our approach with apriori measures or anticipated pathways. Instead our approach has enabled us to gain insights into the psychological process and social patterning of positive change along more typically inductive lines. This is perhaps indicative of the novelty and rapidity of the topic under investigation. However, our analyses do indicate a complex mixture of processes realised within individuals but also across social structures. For example, our sense of the importance of resilience relates to both individual properties but also their position within dynamic social networks.

The strengths of the study lie in its originality and use of a multi-method approach. However, there are some weaknesses that need to be acknowledged. First, some groups, including men, those from lower education groups, and those from BAME communities are under-represented in our survey sample. Due to our purposive sampling approach, the fact that these groups are under-represented may tell us that they were less likely to have experienced positive change. In addition, we sought to overcome this limitation in the sampling approach for the qualitative component whereby we used a sampling matrix in order to recruit a balanced sample in relation to ethnic background, gender, educational attainment, income, and relationship status. A further limitation relates to the retrospective nature of our baseline self-report measures of behaviour which were taken at the same time as our lockdown measures. It is also important to note that the positive changes we report on here were experienced within the context of a national lockdown in Scotland, and the experiences of people living in other national lockdown contexts may have been different, particularly those countries that imposed stricter restrictions on time outdoors.

The current study is the first in the world to take a multi-method, salutogenic approach to COVID-19, and to report on the rapid co-production of research-led resources intended to share insights into initiating positive change. We identified a number of key psychosocial processes that were key to the initiation of positive change, including re-framing, self-reflection and appraisal. In addition, our salutogenic resources that aim to promote positive change represent a starting point from which future salutogenic approaches and interventions may be developed with a focus on building post-pandemic recovery and resilience across people and communities. In particular, targeted interventions are needed to address the inequalities in health that have been amplified by COVID-19, aimed at reducing the social and developmental harms caused by the pandemic.

## Data Availability

Data are not publicly available due to ethical or privacy restrictions

## Acknowledgements

We would like to thank our participants for taking part in the project and our project partners (Actify, Mind the Men, Scottish Older People’s Assembly).

## References

Alonge, O., Sonkarlay, S., Gwaikolo, W., Fahim, C., Cooper, J. L., & Peters, D. H. (2019). Understanding the role of community resilience in addressing the Ebola virus disease epidemic in Liberia: A qualitative study (community resilience in Liberia). Global Health Action, 12(1), 1662682. https://doi.org/10.1080/16549716.2019.1662682

Antonovsky, A. (1979). Health, stress, and coping. San Francisco: Jossey-Bass.

Bauer, G. F., Roy, M., Bakibinga, P., Contu, P., Downe, S., Eriksson, M., Espnes, G. A., Jensen, B. B., Juvinya Canal, D., Lindström, B., Mana, A., Mittelmark, M. B., Morgan, A. R., Pelikan, J. M., Saboga-Nunes, L., Sagy, S., Shorey, S., Vaandrager, L., & Vinje, H. F. (2020). Future directions for the concept of salutogenesis: a position article, Health Promotion International, 35, 187–195, https://doi.org/10.1093/heapro/daz057

Braun, V. & Clarke, V. (2006). Using thematic analysis in psychology. Qualitative Research in Psychology, 3, 77-101. https://doi.org/10.1191/1478088706qp063oa

Cornell, S., Nickel, B., Cvejic, E., Bonner, C., McCaffery, K. J., Ayre, J., Copp, T., Batcup, C., Isautier, J. M. J., Dakin, T. & Dodd, R. H. (2020). What positives can be taken from the COVID-19 pandemic in Australia? medRxiv. https://doi.org/10.1101/2020.12.10.20247346

Ekelund, U.; Steene-Johannessen, J.; Brown, W.J.; Fagerland, M.W.; Owen, N.; Powell, K.E.; Bauman, A.; Lee, I.-M. (2016). Does physical activity attenuate, or even eliminate, the detrimental association of sitting time with mortality? A harmonised meta-analysis of data from more than 1 million men and women. Lancet, 388, 1302–1310. https://doi.org/10.1016/S0140-6736(16)30370-1

Fluharty, M., & Fancourt, D. (2020). How have people been coping during the COVID-19 pandemic? Patterns and predictors of coping strategies amongst 26, 580 adults. PsyArXiv. https://doi.org/10.31234/osf.io/nx7y5

Glaser, B., & Strauss, A. (1967). The discovery of grounded theory: Strategies for qualitative research. Mill Valley, CA: Sociology Press.

Gori, A., Topino, E. & Di Fabio, A. (2020). The protective role of life satisfaction, coping strategies and defense mechanisms on perceived stress due to COVID-19 emergency: A chained mediation model. PloS One, 15, e0242402. https://doi.org/10.1371/journal.pone.0242402

Janssen, X., Fleming, L., Kirk, A., Rollins, L., Young, D., Grealy, M., MacDonald, B., Flowers, P., & Williams, L. (2020). Changes in physical activity, sitting and sleep across the COVID- 19 national lockdown period in Scotland. International Journal of Environmental Research and Public Health, 17(24), 9362. https://doi.org/10.3390/ijerph17249362

Killgore, D. S., Taylor, E. C., Cloonan, S. A., & Dailey, N.S. (2020). Psychological resilience during the COVID-19 lockdown. Psychiatry Research, 291, 113216. https://doi.org/10.1016/j.psychres.2020.113216

Levitt, H. M., Bamberg, M., Creswell, J. W., Frost, D. M., Josselson, R., & Suarez-Orozco, C. (2018). Journal article reporting standards for qualitative primary, qualitative meta- analystic, and mixed methods research in psychology. The APA publications and communications board task force report. American Psychologist, 73, 26–46.

Luthar, S. S., & Cicchetti, D. (2001). The construct of resilience: implications for interventions and social policies. Development and Psychopathology, 12, 857–885. https://doi.org/10.1017/S0954579400004156

O’Connor, R. C., Wetherall, K., Cleare, S., McClelland, H., Melson, A. J., Niedzwiedz, C. L., O’Carroll, R. E., O’Connor, D. B., Platt, S., Scowcroft, E., Watson, B., Zortea, T., Ferguson, E., & Robb, K. A. (2020). Mental health and well-being during the COVID-19 pandemic: longitudinal analyses of adults in the UK COVID-19 Mental Health & Wellbeing study. The British Journal of Psychiatry. https://doi.org/10.1192/bjp.2020.212

Ong, A. D., Bergeman, C. S., Bisconti, T. L., & Wallace, K. A. (2006). Psychological resilience, positive emotions, and successful adaptation to stress in later life. Journal of Personality and Social Psychology, 91, 730–74. https://doi.org/10.1037/0022-3514.91.4.730

Penkler, M., Muller, R., Kenney, M. & Hanson, M. (2020). Back to normal? Building community resilience after COVID-19. The Lancet Diabetes and Endocrinology, 8, 664–665. https://doi.org/10.1016/S2213-8587(20)30237-0

Smith, J. A., Flowers, P. & Larkin, M. (2009). Interpretative phenomenological analysis: Theory, method and research. London: Sage.

Stamatakis, E.; Gale, J.; Bauman, A.; Ekelund, U.; Hamer, M.; Ding, D. (2019). Sitting time, physical activity, and risk of mortality in adults. Journal of the American College of Cardiology, 73, 2062–2072. https://doi.org/10.1016/j.jacc.2019.02.031

Thornley, L., Ball, J., Signal, L., Lawson-Te Aho, K. & Rawson, E. (2015). Building community resilience: learning from the Canterbury earthquakes. New Zealand Journal of Social Sciences Online, 10, 23–35. http://dx.doi.org/10.1080/1177083X.2014.934846

Williams, L., Rollins, L., Young, D., Fleming, L., Grealy, M., Janssen, X., Kirk, A. MacDonald., & Flowers, P. (2020). What have we learned about positive changes experienced during COVID-19 lockdown? Evidence of the social patterning of change. PLoS ONE, 16(10): e0244873. https://doi.org/10.1371/journal.pone.0244873

Wright, L., Steptoe, A., & Fancourt, D. (2020). Are we all in this together? Longitudinal assessment of cumulative adversities by socioeconomic position in the first 3 weeks of lockdown in the UK. Journal of Epidemiology and Community Health. https://doi:10.1136/jech-2020-214475

