## Supplementary figures and images for "Sharing positive changes made during COVID-19 national lockdown: a multi-method co-production study"

### Supplemental File 1

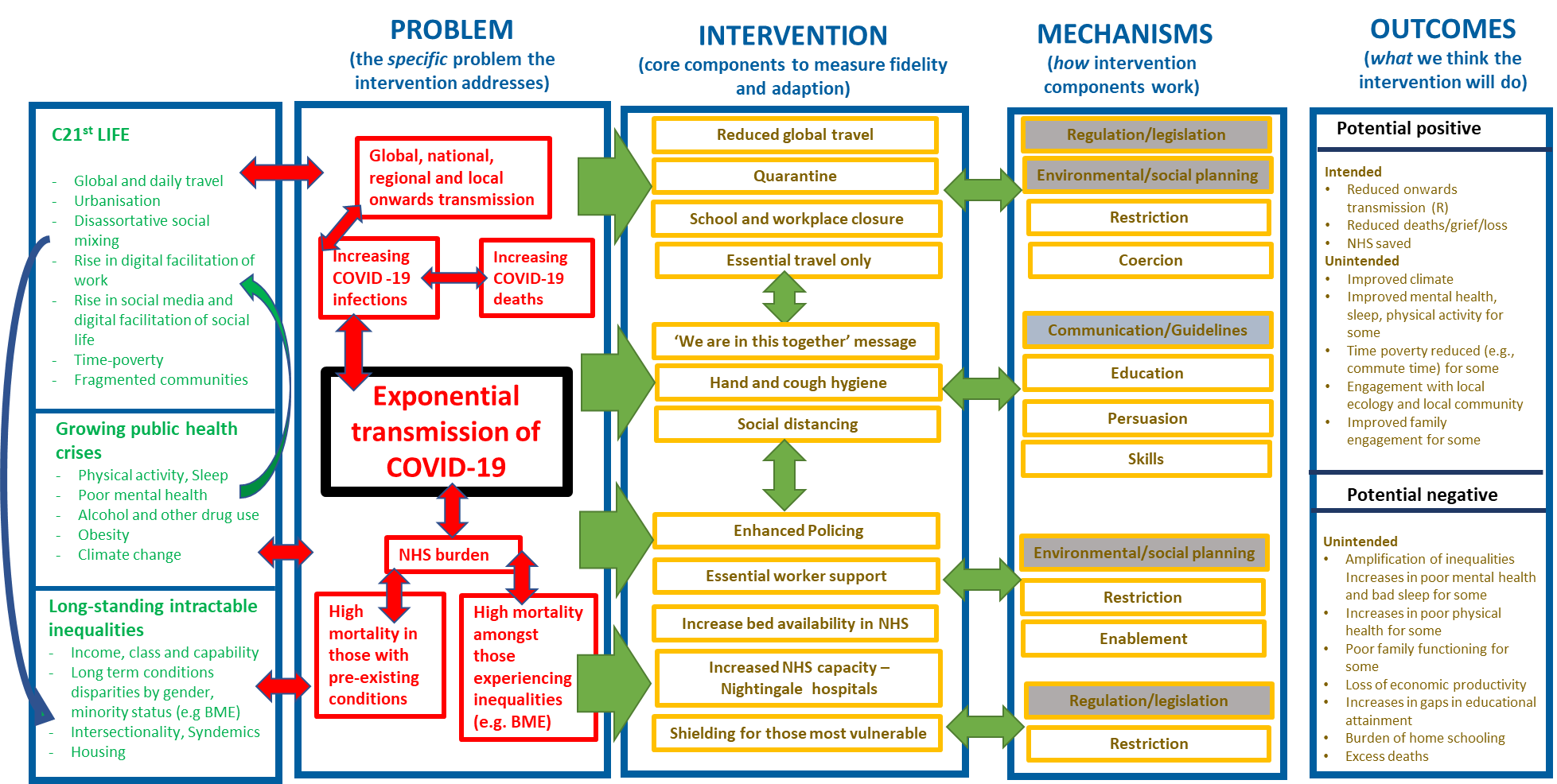
