## Supplemental file 2 for "Sharing positive changes made during COVID-19 national lockdown: a multi-method co-production study"

Supplementary File 2

Distribution of cross-domain positive change scores (%)

| **% change achieved** | **Count** | **Percent** |
| --- | --- | --- |
| 0.000 | 996 | 40.74 |
| 25.000 | 179 | 7.32 |
| 33.333 | 344 | 14.07 |
| 50.000 | 390 | 15.95 |
| 66.667 | 215 | 8.79 |
| 75.000 | 92 | 3.76 |
| 100.000 | 229 | 9.37 |
