## Supplemental file 3 for "Sharing positive changes made during COVID-19 national lockdown: a multi-method co-production study"

**CATALYST Project**

**Top tips for our partners to help us share what we have learned from those who made positive changes during national lockdown**

**Summary**: We talked to 48 diverse people who had experienced positive change during the national lockdown. We analysed how they had experienced change to gain some insights into how they did it so that we could share this with other people and key organisations.

**What we learned from our participants about making positive changes**

| **Top tips for you, and the people you work with, to use to make positive changes in your life and to cope with COVID-19 restrictions** | **Examples of what people said in interviews that you might want to share, use or develop into resources** |
| --- | --- |
| **1. Think about time and what you can do with it** We found people who made positive changes realised that time seemed to have changed and then they often asked themselves what they wanted to do with the time they actually had. Some people had more time than before COVID-19, for example, they got their commuting time back, others had more time because they couldn’t socialise, or because they couldn’t work. Other people had less time because of home-schooling or because they cared for others. Noticing that time had changed was an important step to thinking about making positive changes.  **Helpful questions people can ask themselves**  *How much time do you really have to make changes to your life given new circumstances?*  *What are the things you could do with the time, and circumstances, that COVID has given you?*  *Which are the most important and realistic things that you could achieve in the time you actually have?* | *“And that’s when I just started looking round at different recipes and things like that and trying to mix it up a wee bit. And the government, I think, were giving advice saying, “Try and only go to the shops once. Try and plan what you’re gonna have.” So I Just thought about it. And also, again, that’s another good kinda ‘me’ time to go off into the kitchen for, you know, forty-five minutes by myself, switch the radio on, start cooking,”*  *“It was listening to that, and then realising, oh, actually, rather than listening to music I can listen to audio books. And I found I really, really enjoyed it. And it actually gave me the space, because I wasn’t rushing around doing things, I was just walking from place to place, that I actually could listen to these things and start to reflect on how best to put practices into my life. “*  *“I make quilts… and this is my sewing room. So it’s just – and I suppose again, I don’t know whether it’s right or wrong. It makes you feel needed and gives you a purpose. Not that I’m doing it very quickly, but I can come up here and carry on. Start looking at fabric and get it online, blah, blah, blah, and I suppose that has all developed, and shopping online.”*  *“And so I’ve been cycling. So these things, some of the things which were happening that I had no time to enjoy, or kind of explore, they happened by themselves. So that was one thing. I thought I should improve my music, I said in three months I can do something, you know, concrete, you know, learn something, or learn something new. And it was quite good because my music teacher himself, he thought I could learn certain things, which I would not otherwise have the opportunity to learn. Which needed a lot of concentration and repetition.”*  *“it’s that little pause in time as well, you know, possibly I wouldn’t be here talking to you if it didn’t happen. But, I’m gonna do a… I’m actually going back to uni in August, myself, and when I was… when the lockdown came in I’m thinking, ‘oh should I go and do the course, should I not do the course?’ And, you know, and it’s like, you know, I was having to think a lot about that, whether I was gonna do it or not. And… and then I thought, ‘you know what? If this is not a good time to be studying, during the downturn in the economy, I don’t know when is gonna be a good time to study.’ And it just kinda made me assured that, yes, this is definitely a good way to go for me.”* |
| **2. If you can, make plans and establish a routine** We found that people who managed to make positive changes told us how important planning and routine had been for them. Perhaps because lockdown destroyed many people’s usual routines and daily structures a lot of the people we talked to told us about how hard the first few weeks of the first National lockdown were. People told us about how they managed to make positive changes by actively thinking about time and planning for the medium and long term future, but also about making routines for the present, to get through COVID restrictions day by day. People focussed on making a daily and a weekly routine as well as making plans for the future.  **Helpful suggestions for planning and routine**  *Think about your situation and try and make a routine that you can follow on a daily basis. Try your best to stick to it but don’t be too hard on yourself if you slip up every now and again.*  *Thinking about the time you actually have to make positive changes to your life, think what you could realistically do now, in the next few months or across the next few years. Is there anything in your plans for the next few years that you need to start doing in the next few months or now?*  *Think about your plans and break them down into smaller goals. Start with small easily achievable steps and gradually increase your goals over time but don’t be too hard on yourself if you slip up every now and again.* | *“I decided right at the start of lockdown to make sure that I was, that I had a routine. I don’t know why I decided that, but yeah I decided to get a routine, so daily walks, and also I’ve been threatening to start the gym properly for ages, and I thought, ‘Do you know what? I’m gonna have so much time at home, let’s just do it.’ And I had a pair of dumbbells and a yoga mat and I was like, “Right, that’s it. Let’s start it.” So I started doing exercise five days a week as well!”*  *“All right, one thing I find hard to do is stick to a morning routine. Okay, this guy, this book I’m listening to says ‘start your morning by sitting, you know, by the window and having your coffee, so you get a bit more exposure to light.’ Oh, this works. Oh, this also gives me time to write down in my diary what I want to do today. And it was sort of like this slowly evolving thing of day by day, small bits I could add and then seeing if they worked, and if they didn’t, then moving on to something else”*  *“that’s why I gave myself a weekly plan and also why we roughly stuck to a routine every day as well. Yeah, I’ve got friends who have been, you know, sitting in their pyjamas all day and getting up at twelve and everything, just the thought of that horrified me. I was like, ‘I cannot… I cannot do that’. But also, I think from a mental health point of view because I think, I’ve not always had the best mental health in the past and I thought, I think, ‘I know what I need to do to make sure that I’ve got good mental health.’ And that’s part of it, so…”*  *“Yeah, well, ‘cause people always have goals, but they (cuts out) can be quite vague. So during lockdown when you had more time to think about it, you’re like ‘Well, I’m not’ – I had so much less to do, why not just get some paper out, think about what I want, and then you can start creating and actual path towards that. Rather than just ‘Well, I’d like to do this.’ Like for instance, I’ve always wanted to go to Japan, but I’ve always just thought ‘Oh, I’ll do that at some point.’ But during lockdown I was, like, ‘Well, why not now? Why not get my Japanese books out? Why not look at flights? Why not just do it?’ “* |
| **3. Stop and think about nature and the natural world around you** We found that people who managed to make positive changes to their lives during the lockdown often connected to nature in a new way, or a way that reminded them of their childhood and their perceptions of the wonder of the world. Whilst the human world was changing dramatically because of lockdown, the natural world had its safe and familiar cycles and rhythms. Our participants focused on very different things that reflected their circumstances, including watching the sunrise, watching the leaves on trees change, looking at weeds in the pavement, observing moss growing between bricks, watching the local starlings, growing plants from seeds, watching ducks in the river, seeing shadows and clouds change.  **Helpful things that people can choose to do**  *People can step out of their usual routine and use some time to focus and connect with one or two aspects of the natural world for a while within each of their days.*  *People can try to think about nothing except the living thing that they are looking at.*  *People can try and notice the rhythms and slow changes in the natural world around them.*  *Go to a new local outdoor space and take notice of the wildlife that is present. Take a picture to remember the day.* | *“Although there were anxieties in the background relating to a number of other things, particularly parents, the fact I could just get up and go out and spend some time in nature was…was very positive. And some cases quite exciting. And I was taking a lot of pictures and putting them on Facebook.”*  *“But also just… yeah, there’s a safety in nature, I find, and just, I don’t know, a comfort. Like I’m really comfortable being in soil and like smelling grass. Yeah. (And…) I’m noticing… and that’s the other thing I’ve noticed, like I’ve, I don’t think I’ve gone through a Spring or a Summer where I’ve smelt so many different floral scents as you’re walking past trees and, you know, bits, green bits. I’m like, ‘oh that’s a new smell,’ so like that’s really struck me as well, and like that discovery is quite joy inducing. (Laughter)”*  *“But I guess particularly, you know, up along, you know, sort of the canal just, you know, sort of seeing swans and birds and you know, if we are out with friends we’d recognise things – “Oh, that’s a such-and-such” and, you know, it was just really nice and just not realising that was in the heart of Maryhill or, you know, the heart of industrial Glasgow that, you know, there was such nice areas around. So it was just – you appreciate it more I guess, you also are just finding things to distract yourself with and I guess nature provides lots of distraction like that. “*  *And looking at the view, looking at the animals and the birds, of which there are an awful lot more round here than we’ve experienced elsewhere, I think, really.*  *“We’ve been rediscovering simple pleasures like watching the blue tits in the bird box. (Laugh) or a really obvious, not obvious, but vivid example. We have rhubarb in the garden. It just grows there. For the last twenty years, but this year, I have noticed it unfurling its leaves more. And, you know, I wouldn’t normally, I would only look at it at the weekend maybe when I went into the garden. So it’s something, I think, we, at least my family and I think others are becoming more attuned to nature and our immediate environment. Even if it’s just one tree you can see out the window that we notice the gradual changes*  *“I think it, it kinda started because I realised that I... I needed to do an activity that could take up a lot of time, but at... but at the same time, you know, kinda like helped me to practice, you know, patience and things like that and... well, essentially gardening’s that, ‘cause from the start of it, you know, you start off with a seedling and, you know, you do have to keep... keep gardening. Well you do have to keep visiting the plant for like multiple times in a day, you know, you’ve got to go and check on it, and then you’ve got to water it, and then you’ve got to switch position in the house if there’s no sun, and I think that kinda like, that makes you reflect about how this whole lockdown journey has been.”* |
| **4. Reach out for support** We found that those people who made the most positive change reached out to others to help them with their changes. People’s situations were very different and they sought support from all kinds of sources. These included friends and family, online platforms and social media.  **Helpful things that people can choose to do**  *If you can, be proactive, in seeking help and support. Most people will be pleased to provide it.*  *If you can, access support in more than one way, so use social media as well as your friends and family.*  *If it feels safe, share your plans and ideas with other people, particularly if they might share some of the same plans.* | *“Yeah, so, and… and also it seemed a few people online like friends and other people I know who are into gym stuff and just asking them and, do you know what they all—they all said, “Just start. Just do it.” Like, “Don’t take too long thinking about just get on and do it.” So, yeah, I’ve kinda taken that on board…”Yeah, friend group and, you know, people that you follow online and stuff as well. Just messaging them and saying, “Oh, I see you were doing this. How would I go about doing… kinda getting started.”*  *“I Skype them every day until May….just to have someone there I could talk to, ‘cause I didn’t feel like a burden to my parents. So that was really nice and now we’ve weaned down, my mum is back to work so we’ve, we weaned down to two or three times a week in May and now once a week in June ‘cause they’re both back to work. And so that’s been really nice to just also, ‘cause obviously I won’t see them this year ‘cause we had to cancel our trip home. And so it’s been, it’s been nice and I think that’s continued. I’m really close to my parents so obviously wasn’t a big change. But it was nice to have their presence.”*  *“The main one that’s made a tremendous difference, my daughter-in-law’s the yoga teacher but she has done yoga sessions Tuesday, Thursday, Friday and a Sunday morning. And that’s just been a lifeline, because you can have a chat with people who are in your class before and after. Not that great deal of time but at least it’s human contact out-with the house.”*  *“But I’ve also been able to do online ballet classes every day. I’ve been able to link up with this choir that I was with. The people running the choir took steps to… to go online, so they worked very hard to develop an online provision for the members.”*  *“I wasn’t sure what to do. But I did find a programme on YouTube, and it was a lady that was walking. So it was walking, cardio, and weight. So I could walk, do a walk for two or three miles a day and come in and do an hour with this lady on YouTube. And because I thought that if corona is like legionella then if it’s inhaled that if I get fitter and it would change my breathing. So I would’ve… it would change my lung capacity. So that was what was in my head.”*  *“I think what’s happened is, people have become very isolated in themselves. Everybody’s scared, so they have their own worries, and they don’t really think about other people. Whereas I think what I’ve done is, I’ve taken control of our situation, and then tried to help other people. And I think from what I’ve read, you know, various information that’s being sent out, that those people who help others are actually the people that have found lockdown a lot easier, than those people who have just focussed on themselves. And their fears and worries.”* |
| **5. Try connecting with neighbours and your community.** Through doing the interviews, we found that many people who made positive changes connected to their community to help keep them busy and also feel a sense of connection to the community that they felt they were missing before lockdown. This helped them build important social connections that did not exist before lockdown and also gave them a sense of purpose. This took many forms, including starting WhatsApp groups with neighbours, speaking over fences, sharing gardening techniques, picking up medications and essential items for other people.  **Helpful things that people can choose to do**  *Ask vulnerable neighbours if they need anything from the shops. If you feel it is safe to, give them your phone number if you are willing to help them out on other occasions.*  *Share extra resources you have with other members of your neighbourhood. A small gift is a great way to start a friendship. People told us that these included: off cuts of plants, books, baking, garden access for children etc.*  *Start a WhatsApp group with the neighbourhood. This can be an easy casual way to get to know each other without it being publicly online like on other platforms.* | *“the accidental germination of the seeds happened before lockdown. But I think where this actually started becoming positive is that we have a neighbourhood WhatsApp group. So it’s kinda for people who… ‘cause there is quite a few elderly people who live in my neighbourhood, and a couple of pregnant women as well, who are very vulnerable. So we’re all kinda looking out for each other a little bit. But I remember one day just posting on that- on that group, saying, ‘help, I’ve got sixty pepper plants, somebody please, please, please, would you like one?’ And loads of people did, so I literally… I was mid-summer Santa, you know, I just put all the plants in a tray and just went around the different houses and just handed them out.”*  *“And I would actually also say that I now know a lot more o’ my neighbours, a lot better than I ever did before, because I used to sorta wave to them or I’d speak to ‘em and just say “Good morning” to them on the way past, but now we actually have 20 minute discussion, you know? Socially distanced, of course, you know, where we stand – we stand a good bit away and we have a good blether now for a long time now. We know far more about each other now than we ever did before..You know?, so – so I’m actually far closer now to people, to my neighbours and friends here than I ever was before, you know?”*  *“Lots of flowers and vegetables for my own – for my own self, and I’ve also been giving them to a lot of my neighbours. I have some elderly neighbours here who stay residential. I’ve been giving them flowers and vegetables and everything I’ve been growing.”*  *You’re, you know, you perhaps see each other if you’re doing a bit of gardening or anything. So we’ve got really, really close to our neighbours*  *“There’s a lot of people mental health issues, emotional problems, and, of course, you’ve got refugees and asylum seekers as well. And I think because of the lockdown, there’s been a more sympathetic attitude, I think, towards a lot of people wi’ problems like this and I think going forward, it would be great if society could keep that up. And be a bit more “every person counts” kinda attitude rather than sort of “me, me, me” kinda thing”* |
| **6. Incorporate some physical activity into your days to support your mental wellbeing.** We found that many of the people we talked to turned to exercise and physical activity to reduce stress in their lives, and clear their heads while going through this difficult time. The type of activity and amount varied, but people told us that they felt mentally stronger after taking up some physical activity. There were many examples of activities used including indoor activities supported by online platforms like YouTube and zoom, discovering new local areas on walks and meditation practices like yoga.  **Helpful things that people can do**  *Start small with your goals and try not to set unrealistic expectations.*  *Test the waters before you sign up or commit to online exercise classes. It is ok to change what you want to do.*  *Take some time to research online what type of activity you might enjoy. There are many options for both indoor physical activity and outdoor physical activity.*  *Ask friends or family what physical activities they turn to when they are feeling stressed and need a break.*  *It does not to be a large amount! 10 minutes of walking in the fresh air is often enough to clear your head.* | *“Okay, so, yeah, so if I’m honest, I found it really difficult in the beginning and I felt my mental health was declining because I wasn’t getting out of the house and I was missing the interaction with colleagues. And I missed the work, you know, I go to a lot of meetings at work..And I think it would’ve been… I think, you know, I think that’s why we shifted from the baking thing that we need to do an outdoor activity because I was beginning to feel a bit down because, you know, when it started I thought, ‘Oh, this will only be a week or two…’ (laugh) naively. You know, and I kept thinking—but, you know, as the kind of days went into weeks, and I thought, ‘You know what? This isn’t good.’ And, as I say, my sleep started to be affected. And that really made me feel quite down. So that’s where the walking really helped, very quickly I felt the mood was lifting by doing that and it was something to look forward to. And, actually, just getting the fresh air and, do you know what? I really enjoyed just discovering where we live. You know, different places around, different parks, you know, some of the lovely walks and actually, that was good. But the biggest impact, I think, was just feeling better in myself and feeling fitter and healthier.”*  *“And back, so I’m not someone who ever thought about exercise. I’ve never been to a gym, you know, I don’t go out and do exercise for that reason. I mean I’ll go out for a walk, ‘cause I like going for a walk. And what I noticed a few weeks into lockdown was I was turning into a couch potato because I no longer had exercise built into my routine, in my commute. So that was quite a thing for me to get my head round that I would now have to be, I would now have to think, ‘I must exercise and do something.’ And that took quite a while for me to get my head round, but I’m sure that is a positive, you know, in the long term, I’m, what am I? Fifty-something now, that is something I’ll have to get used to. I’m too—when I was young, I could get away with not exercising, but now I’m getting older, menopause and all that, it makes—you have to think about exercise.”*  *“yeah. I feel like it is literally to just be able to... to get some fresh air. And there have been times as well like during lockdown I’ve had like some stressful stuff or an interview or something, and I have gone out for a walk beforehand to think things through like just myself, and just... and hope that it’ll just clear my mind a little bit. So I’ve done that a little bit as well, sort of as a de-stressor.”*  *“And I suppose there was a conscious effort in terms of my own mental health where I had to take seriously my physical wellbeing, with the closure of the gyms. That had been my go-to, my headspace, if you like, and that was being taken away from me as well because the gyms were closing. So I had to put in a strategy, so I done running. And I don’t like to run. It’s not my favourite thing in the world, but it was something that I could do in the hour that we got, the hour of freedom, to keep that headspace. So keeping that up was important. There was a lot of motivators behind that in terms of my physical and mental health. So that was a positive change is the running.”* |
