## Supplemental file 4 for "Sharing positive changes made during COVID-19 national lockdown: a multi-method co-production study"

Supplementary file 4

*Univariate analysis of socio-demographic and psychological factors predicting ≥50% positive change*

| ***Variable*** | ***p-value*** | ***Comparison*** | ***Coefficient*** | ***p-value*** |
| --- | --- | --- | --- | --- |
| Age | <.001 | 18-24 vs. 65+ | 1.07 | .728 |
|  |  | 25-34 vs. 65+ | 1.52 | .010 |
|  |  | 35-49 vs. 65+ | 2.01 | <.001 |
|  |  | 50-64 vs. 65+ | 1.73 | <.001 |
| Gender | .002 | Female vs. Male | 1.41 | - |
| Relationship status | .040 | Married/living with partner vs. Single | 1.30 | .019 |
|  |  | Have a partner but not living together vs. Single | .969 | .864 |
|  |  | Separated/divorced/widowed vs. Single | 1.080 | .673 |
| Ethnicity | 0.105 | White vs. BAME | 1.58 | - |
| Education | 0.506 | High School vs. Postgraduate | .810 | .148 |
|  |  | College vs. Postgraduate | .927 | .550 |
|  |  | Undergraduate vs Postgraduate | .989 | .908 |
| Household income | .072 | <£16, 000 vs £90,000+ | .674 | .042 |
|  |  | £16,000-29999 vs £90,000 | .701 | .036 |
|  |  | £30,000-£59999 vs £90,000 | .877 | .389 |
|  |  | £60000-£89999 vs £90,000 | .924 | .639 |
| Employment status | <.001 | Inactive vs. Employed | .671 | <.001 |
|  |  | Unemployed vs. Employed | .694 | .304 |
| Overall health | .012 | Very poor vs very good | .284 | .252 |
|  |  | Poor vs Very Good | .761 | .404 |
|  |  | Fair vs Very Good | .635 | .001 |
|  |  | Good vs Very Good | .851 | .076 |
| High risk/Shielding | <.001 | Yes vs. No | .613 | <.001 |
| Social support | <.001 | - | 1.34 | - |
| Anxiety | <.001 | - | .842 | - |
| Depression | <.001 | - | .785 | - |
| Positive reframing | <.001 | - | 1.42 | - |
| Acceptance | .001 | - | 1.20 | - |
| Planning | .002 | - | 1.16 | - |
| Humour | .048 | - | 1.08 | - |
| Active coping | <.001 | - | 1.25 | - |
| Emotional support | .003 | - | 1.15 | - |
| Self-distraction | .001 | - | .845 | - |
| Instrumental support | .006 | - | 1.17 | - |
| Personal competency | .001 | - | 1.35 | - |
| Social resources | .001 |  | 1.28 | - |
